# Young Zambian infants with symptomatic RSV and pertussis infections are frequently prescribed inappropriate antibiotics

**DOI:** 10.1101/2021.09.21.21263905

**Authors:** CE Gunning, P Rohani, L Mwananyanda, G Kwenda, Z Mupila, CJ Gill

**Affiliations:** University of Georgia, Odum School of Ecology; Boston University School of Public Health, Department of Global Health; Right to Care – Zambia; University of Zambia, School of Health Sciences, Department of Biomedical Science; University of Georgia, Center for the Ecology of Infectious Diseases; University of Georgia, Department of Infectious Diseases

## Abstract

Pediatric community-acquired pneumonia (CAP) remains a pressing global health concern, particulary in low-resource settings where diagnosis and treatment rely on empiric, symptoms-based guidelines such as the WHO’s Integrated Management of Childhood Illness (IMCI). This study details the delivery of IMCI-based health care in a cohort of 1,320 young infants and their mothers in a low-resource urban community in Lusaka, Zambia. We prospectively monitored mother/infant pairs across infants’ first four months of life, recording symptoms of respiratory infection and antibiotics prescriptions (predominantly penicillins), and tested nasopharyngeal (NP) samples for Respiratory syncytial virus (RSV) and *Bordetella pertussis*. Symptoms and antibiotics use were more common in infants (43% and 15.7%) than in mothers (16.6% and 8%), while RSV and *B. pertussis* were observed at similar rates in infants (2.7% and 32.5%) and mothers (2% and 35.5%), albeit frequently at very low levels. In infants, we observed strong associations between symptoms, pathogen detection, and antibiotics use. Critically, we demonstrate that non-macrolide antibiotics were commonly prescribed for pertussis infections, some of which persisted across many weeks. We speculate that improved diagnostic specificity and/or clinician education paired with timely, appropriate treatment of pertussis could substantially reduce the burden of this disease while reducing the off-target use of penicillins.

## Introduction

Pediatric community-acquired pneumonia (CAP) remains a pressing global health concern, accounting for nearly 20% of deaths in young children [1, 2]. Yet appropriate diagnosis and treatment of CAP remains challenging: viral CAP is common in children [3], while viral and bacterial CAP are difficult to distinguish [4–6]. Quantitative PCR (qPCR) allows for rapid, specific, and relatively inexpensive identification of certain pathogens [3, 7, 8], but timely diagnostic capacity is not widely available in low-resource settings. In order to improve child healthcare outcomes globally, the World Health Organization’s Integrated Management of Childhood Illness (IMCI) was established in the mid-1990s and has since been widely adopted and periodically updated [9–12]. The modern IMCI provides empiric, symptoms-based diagnosis and antibiotics-based treatment of pediatric CAP that are inexpensive and widely available. Of note, IMCI guidelines are biased towards sensitivity and over specificity, as the immediate cost of inaction is potentially high.

Respiratory syncytial virus (RSV) and *Bordetella pertussis* are two common childhood pathogens that cause widespread and sometimes severe disease, particularly in low- and middle-income countries (LMICs) [2, 13]. However, both RSV and pertussis share symptoms with a range of other childhood diseases, complicating surveillance and diagnosis. RSV remains a leading cause of pediatric CAP worldwide, with morbidity and mortality overwhelmingly concentrated in the very young and in LMICs [13, 14]. Widespread pertussis vaccination has dramatically reduced infant mortality worldwide [15, 16], though older and more reactogenic whole-cell vaccines (wP) remain the standard of care in LMICs [17, 18]. Unlike acellular pertussis vaccines (aP), however, wP booster doses cannot be administered to pregnant mothers during prenatal care due to reactogenicity [18]. For RSV, no licensed vaccine or chemical treatment exists, though recent advances in maternal vaccination and long-lasting monoclonal antibodies show promise as prophylactic interventions [19, 20].

Pediatric RSV infections are typically short-lived (approximately 1-2 weeks) but do not provide durable immunity, and repeat infections are common [14, 21]. Pertussis infections can last much longer (2-8 weeks of acute respiratory symptoms) but can be effectively treated with macrolide antibiotics [22, 23], though macrolide resistance is a growing concern throughout East Asia [24, 25]. *B. pertussis* infections are thought to confer long-lasting immunity, with considerable debate surrounding the duration of both natural and vaccine-derived immunity [17, 26–28]. For both diseases, repeat infections are less severe, and the symptom severity generally decreases with age, though RSV’s impact on the elderly remains a concern [29]. And while morbidity and mortality in otherwise healthy adults is limited, the epidemiological impact of infections in adults, particularly caregivers, on transmission to children remains an open question [30, 31].

Critically, neither RSV nor *B. pertussis* are directly targeted by IMCI guidelines, where treatment of uncomplicated CAP (cough or difficulty breathing and fast breathing or chest indrawing) includes a 3-day course of oral amoxicillin [1]. Amoxicillin is a preferred front-line antibiotic due to its low cost, ease of administration, low side-effect profile, and broad efficacy [1, 32, 33], yet it (and penicillins in general) is not a recommended treatment for pertussis [23], even though direct evidence for its efficacy against *B. pertussis* is limited and possibly outdated [34].

In this study, we examine pediatric and maternal health care delivery in a low-resource urban setting where IMCI guidelines direct empiric clinical treatment of respiratory infections. Using prospective longitudinal surveillance of a cohort of 1,320 young infants and their mothers, we employ an epidemiological approach to assess the interdependence between RSV and *B. pertussis*, symptoms of respiratory infections, and antibiotics use in mother/infant pairs. Our goal is to quantify A) the frequency of symptomatic RSV and pertussis infections, and B) the associated frequency of non-macrolide antibiotic use to treat these two off-target pathogens. Further, we seek to elucidate the clinical presentation and treatment of RSV and pertussis in this population, and to compare disease presentation between infants and their mothers, whose immune histories greatly differ.

## Results

### Overview

During 2015, we recruited mothers and their healthy newborn infants (aged 0-10 days) from Chawama township, a densely-populated urban community in Lusaka, Zambia. We prospectively monitored subjects across infants’ first four months of life during visits to the sole public health clinic (PHC) in this community. At each of seven scheduled study visits (and additional mother-initiated visits), subjects received routine, no-cost health care from PHC clinicians who followed IMCI guidelines for symptoms-based diagnosis and treatment of acute infections. In addition, we collected nasopharyngeal (NP) samples (including enrollment visit) and recorded symptoms of respiratory infection and whether antibiotics were prescribed (excluding enrollment visit). NP samples were retrospectively tested for RSV and *B. pertussis* using qPCR.

Study details, including enrollment criteria, study profile, and demographics, are provided in Gill et al. [35] and Gill et al. [36], and the timing of wP dose administration in this cohort is detailed in Gunning et al. [37]. In this cohort we have previously described ten cases of clinical pertussis in infants [35], as well as evidence for more frequent mild and asymptomatic pertussis in both mothers and infants [36].

We focus here on the analysis set used in Gill et al. [36]: the 1,320 mother/infant pairs with ≥ 4 NP samples per subject. To better evaluate routine clinical care in a representative low-resource setting, we exclude unscheduled, mother-initiated visits from the analysis set (unlike Gill et al. [36]), for a total of 8390 visits and 16784 NP samples. We categorize any detectable qPCR signal, i.e., CT≤45, as a detection of RSV or pertussis in a NP sample, since our goal is population-level surveillance rather than diagnosis of disease in individuals. We note that qPCR testing of NP samples was retrospective, and that these results were not available to clinicians as part patient care.

The overall frequency of outcomes are shown in Table 1, stratified by unique subjects and visits (columns) where the outcome was observed: antibiotics prescription during visit (ABX), self-reported symptoms of respiratory infection, and detection of pertussis and/or RSV. In Figure 1, we illustrate the co-occurrence of all pairs of outcomes within study visits, stratified by mothers versus infants (Figure 1A), or the co-occurrence of each outcome during visits by mothers and their infants (Figure 1B).

**Table 1:** Frequency (and percent) of unique subjects and visits where each study outcome was observed, out of 1320 mother/infant pairs and 8390 regularly scheduled clinic visits (including enrollment). In addition, six infants in the analysis set died during the study. ABX: antibiotics prescribed during visit. Symptoms: symptoms of respiratory infection.

| Outcome | Subjects | Visits |
| --- | --- | --- |
| **Infants** |  |  |
| ABX | 207 (15.7) | 258 (3.1) |
| Symptoms | 568 (43.0) | 768 (9.2) |
| Pertussis | 429 (32.5) | 670 (8.0) |
| RSV | 36 (2.7) | 41 (0.5) |
| Pertussis & RSV | 6 (0.5) | 6 (0.1) |
| **Mothers** |  |  |
| ABX | 106 (8.0) | 124 (1.5) |
| Symptoms | 219 (16.6) | 240 (2.9) |
| Pertussis | 471 (35.7) | 759 (9.0) |
| RSV | 27 (2.0) | 28 (0.3) |
| Pertussis & RSV | 4 (0.3) | 4 (0.0) |

**Figure 1.**
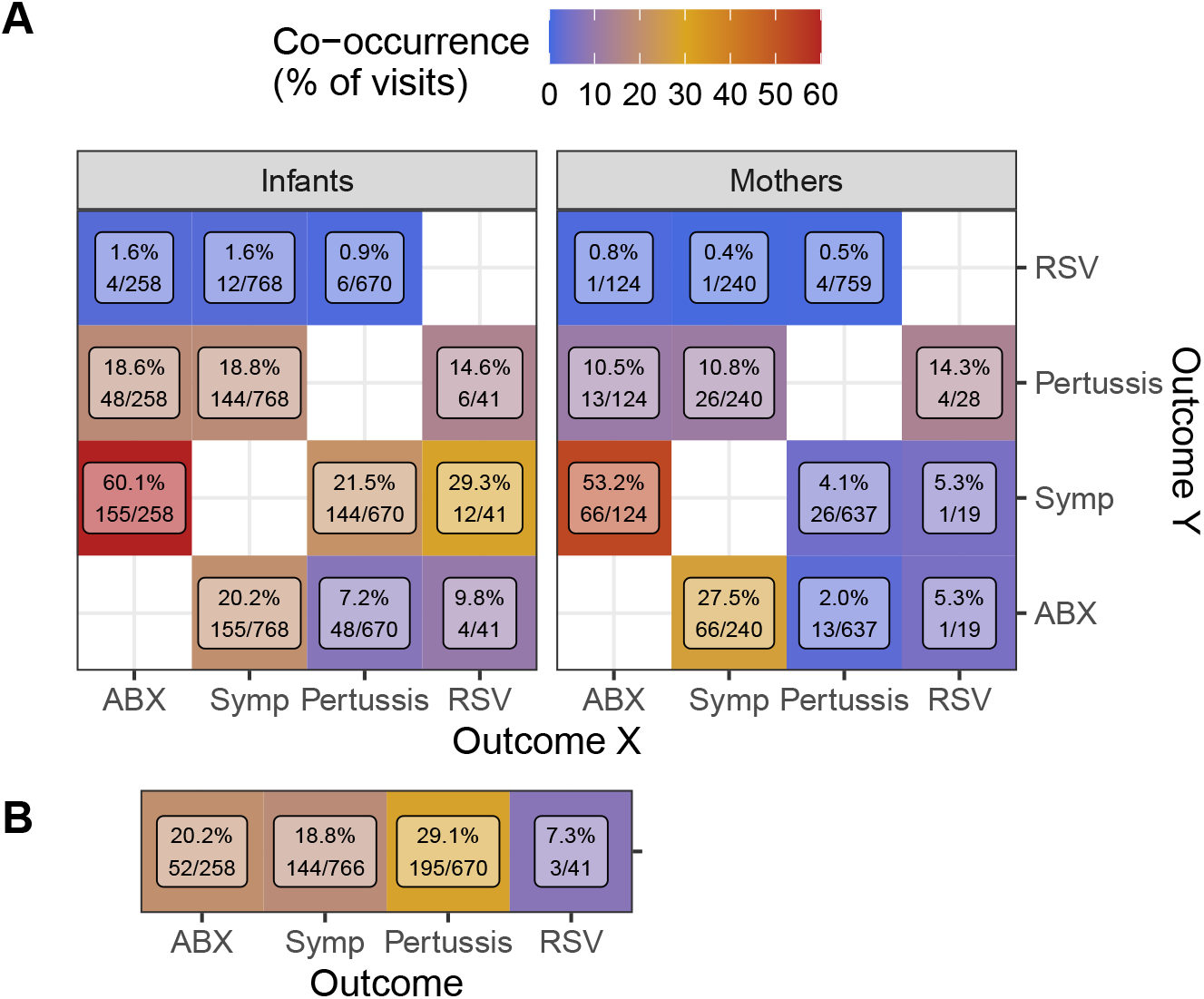
Co-occurrence of outcomes (rows, columns) during study visits. **A)** Within infants (left) or mothers (right), percent of visits with outcome X (column) in which outcome Y (row) was also observed (Y/X). **B)** Percent of infant visits with each outcome (columns) where the outcome was also observed in the infant’s mother (Mother/Infant). Text shows percent co-occurrence and the number of visits in the numerator and denominator. Symptoms (Symp) and prescriptions of antibiotics (ABX) were not recorded at enrollment visits, but only apparently healthy infants were enrolled.

Overall, we observe the highest co-occurrence between antibiotics use and symptoms (infants=60.1% of visits with antibiotics prescriptions, mothers=53.2%), and between RSV detections and symptoms in infants (29.3% of visits with RSV detections). In addition, subjects were commonly prescribed antibiotics during visits where symptoms were recorded (infants=20.2% of symptomatic visits, mothers=27.5%). We also find a substantial co-occurrence between infants and their mothers for antibiotics use (20.2% of infant visits with antibiotics prescriptions), symptoms (18.8%), and pertussis detections (29.1%).

In addition to descriptive statistics, we also use a set of logistic generalized linear models (GLMs) to assess the presumed causal relationship between pathogens (RSV and pertussis), symptoms, and antibiotics use. Our presentation assumes that pathogens cause symptoms, and that antibiotics were prescribed in response to symptoms.

### Prevalence of RSV and pertussis

In Figure 2, we show the estimated proportion of NP samples where RSV (Figure 2A) or pertussis (Figure 2B) was detected across the study period. We also show NP sampling intensity over time (Figure 2C), which reflects the cohort’s rolling enrollment. In March 2015, at the beginning of our study, we observe a brief period of elevated RSV prevalence that suggested the end of a seasonal outbreak, consistent with previous observations in Lusaka and similar locales [38, 39]. This was followed by an outbreak of pertussis that stretched from May to August 2015, during the coolest months of the year in Lusaka. We detected pertussis in 32.5% of infants and 35.7% of mothers in this cohort, and in 8% and 9% of infants’ and mothers’ NP samples, respectively (see also Gill et al. [36]). Multiple pertussis detections were common: we observed ≥2 detections in 12% of infants and 13.9% of mothers, and ≥3 detections in 4.5% of infants and 5.7% of mothers. We detected RSV in 2.7% of infants and 2% of mothers, though repeat detections were rare (3 infants and 1 mother). Both RSV and pertussis were simultaneously detected in just 6 infants and 4 mothers (0.5% and 0.3%, respectively).

**Figure 2.**
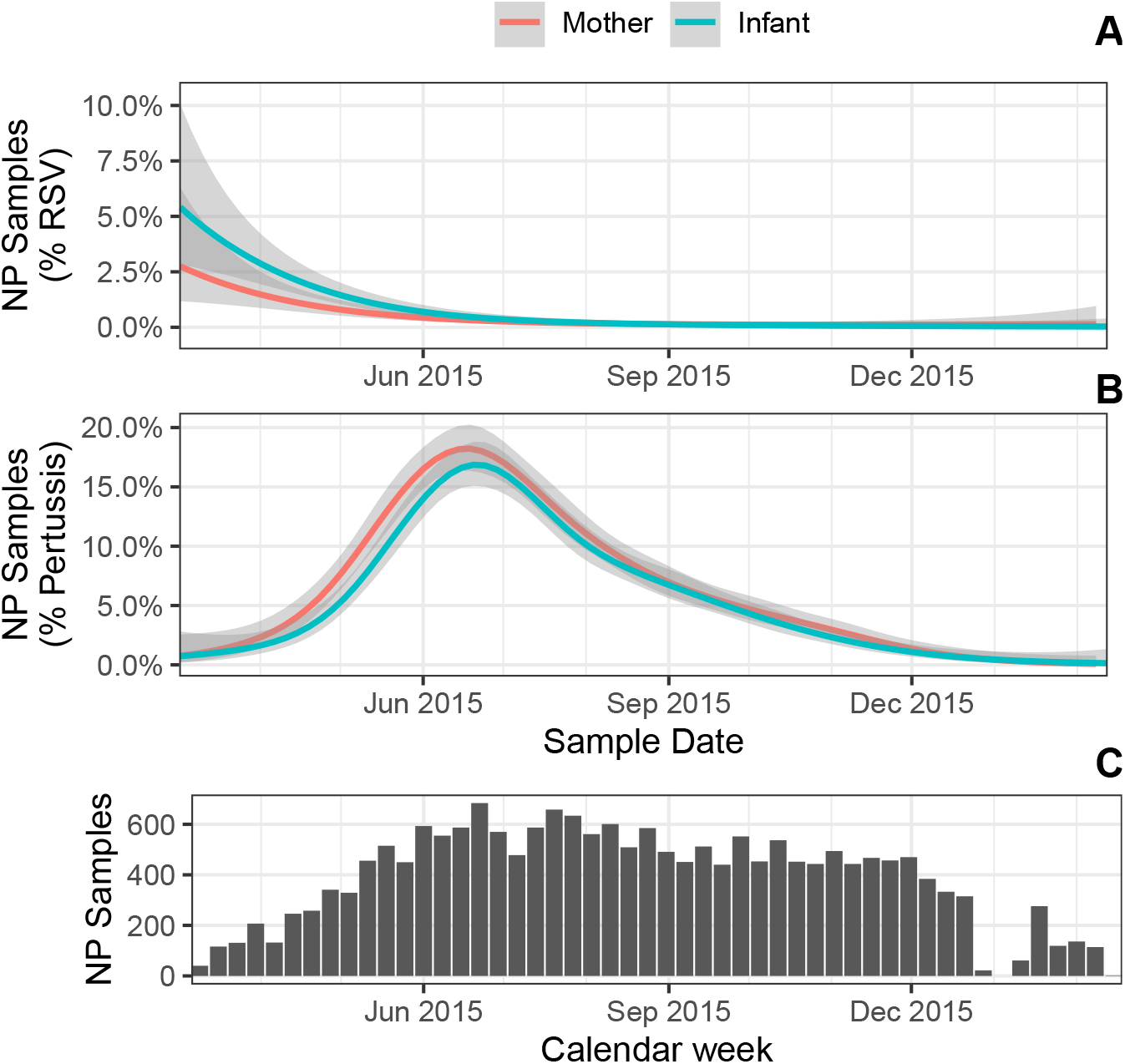
**A-B)** percent of NP samples with detectable RSV (A) or IS481 (B) over time. Shading shows 95% CI (estimated by GAM); color indicates mothers versus infants. **C)** NP samples per calendar week (N=17,442).

### Symptoms of respiratory infections

Symptoms were reported and/or observed in 568 infants (43%) across 768 visits (9.2%), and in 219 mothers (16.6%) across 240 visits (2.9%) (Table 1). Most symptoms were mild: simple cough and/or coryza alone accounted for 84.9% (infants) and 90.8% (mothers) of visits where symptoms were recorded. In addition, simple cough and/or coryza were sensitive indicators of other symptoms, such that only 3.5% (infants) and 5% (mothers) of visits where any symptoms were recorded included neither cough nor coryza.

As expected, visits with symptomatic pertussis detections were much more common in infants (21.5%) than in mothers (4.1%) (Figure 1). As previously described [35], severe pertussis was rare in this cohort. And, while RSV detections were rare overall, symptomatic RSV detections were much more frequent in infants than mothers (12/41 visits and 1/19 visits, respectively).

Logistic regression showed that an infants’ odds of symptoms were approximately 3-fold higher when pertussis was detected, and 4.7-fold higher when RSV was detected (Table 3). We also observed a modest increase in the odds of symptoms as infants age (OR=1.06 per week of age). We presume this increase corresponds with infants acquiring infectious diseases across the course of the study, consistent with our previous findings of age-dependent pertussis prevalence in study infants [36]. Also consistent with our previous findings, we do not observe a consistent relationship between RSV or pertussis and symptoms in mothers.

Most pertussis detections in this cohort exhibited weak qPCR signals that would be considered ‘negative’ in clinical settings (CT>40) [36]. We did, however, observe modestly lower IS481 CT values when symptoms were present versus absent in infants (median 41.8 vs 43.4, p<0.001) but not in mothers (Figure 3). We found a similar pattern in RSV (0.9 difference of medians in infants), albeit with very low sample sizes and weak strength of evidence (*p*=0.3).

**Figure 3.**
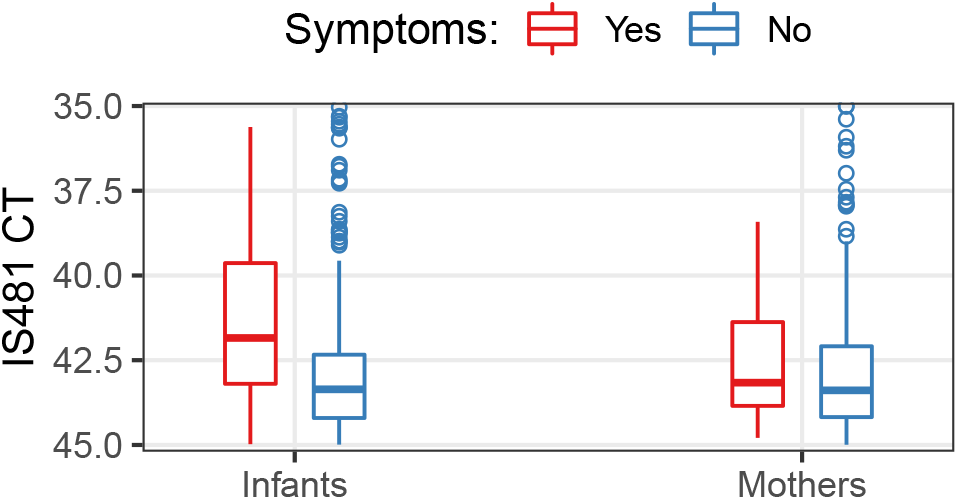
Distribution of IS481 CT values: boxplot stratified by the presence of symptoms (color). In infants, the median value was 1.35 higher when symptoms were present (95% CI: 1.0-1.7). For clarity, NP samples with CT<35 are not shown (20 out of 16,784 total NP samples).

### Antibiotics use

Overall, we recorded 382 antibiotics prescriptions for 313 unique subjects. Approximately twice as many infants received an antibiotics prescription during the study as mothers (207 infants versus 106 mothers). Infants also received more antibiotics than mothers (Table 2): while a single mother received 3 prescriptions, ten infants received ≥ 3 prescriptions (max 4).

The recorded duration of most prescriptions was five days (79.8%), followed by seven days (10.2%), and not recorded (6%). Most prescriptions were for penicillins (62% and 67% in infants and mothers), followed by cephalosporins (17.1% and 3.2%), undetermined (11.6% and 21%), along with infrequent prescriptions of aminoglycosides or nitroimidazoles (2.3% and 6.5% in infants and mothers). Use of macrolides and sulfonamides were rare, accounting for 6.2% and 2.4% (macrolides) and 3.1% and 3.2% (sulfonamides) of prescriptions in infants and mothers, respectively.

**Table 2:** Number of unique subjects that received antibiotics, stratified by the number of clinic visits at which antibiotics were prescribed (rows).

| Visits | Infants | Mothers |
| --- | --- | --- |
| 1 | 167 | 89 |
| 2 | 30 | 16 |
| 3 | 9 | 1 |
| 4 | 1 | 0 |
| Sum | 207 | 106 |

In Table 4, we detail how antibiotics use in infants and mothers changed between clinic visits, and in response to pathogens. We observed a large positive association with antibiotics prescription at the previous clinic visit (infant OR=5.4, mother OR=6.4, p<0.001), and a high correspondence of antibiotics prescriptions within mother/infant pairs (infant OR=19.7 and mother OR=25.2, p<0.001). Albeit with weak statistical evidence due to small sample sizes, we observe that RSV detections predicted antibiotics use in infants (OR=3.2, *p*=0.055). Interestingly, we observe that contemporaneous pertussis detection corresponded with increased antibiotics use in infants (OR=2.2, p<0.001) but not mothers. Conversely, detection of pertussis at the previous visit predicted antibiotics use in mothers (OR=2.3, *p*=0.004), with a less pronounced association in infants (OR=1.5, *p*=0.047).

**Table 3:** Symptoms of respiratory infection: estimated effects of covariates on the presence of symptoms. Separate GLMs were constructed for infants (N=8390 visits) and mothers (N=7074 visits)

| Covariate | OR | P | 95% CI |
| --- | --- | --- | --- |
| **Infants** |  |  |  |
| Infant Age (wks) | 1.06 | <0.001 | 1.04 - 1.08 |
| Pertussis | 2.95 | <0.001 | 2.40 - 3.61 |
| RSV | 4.69 | <0.001 | 2.23 - 9.25 |
| **Mothers** |  |  |  |
| Infant Age (wks) | 0.96 | 0.022 | 0.93 - 0.99 |
| Pertussis | 1.25 | 0.3 | 0.81 - 1.86 |
| RSV | 1.53 | 0.7 | 0.08 - 7.45 |

**Table 4:**
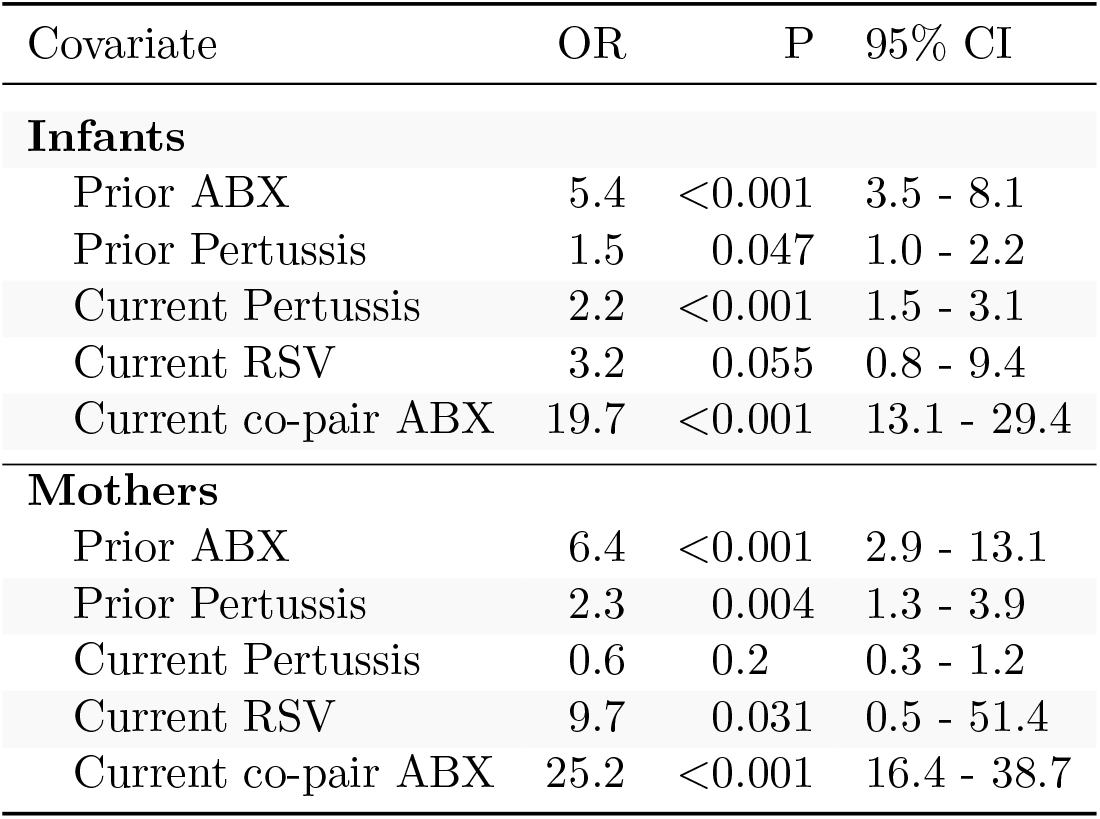
Prescription of Antibiotics: estimated effect of covariates at prior or current visit, relative to prescription. OR: odds ratio. Separate GLMs were constructed for infants (N=7066 visits) and mothers (N=5748 visits). Co-pair indicates the status of an infant’s mother and a mother’s infant, respectively.

We also considered whether prescription of antibiotics at the previous clinic visit was associated with subsequent detection of pertussis. To test this association, we only considered antibiotics use *prior* to a subject’s first detection of pertussis, since we found that antibiotics were commonly prescribed in *response* to pertussis. Consistent with modern clinical guidelines, we do not find compelling evidence that antibiotics use affected pertussis detection at the subsequent visit (infant OR=1.3, *p*=0.3 and mother OR=0.7, *p*=0.4).

For illustrative purposes, we display in Figure 4 the study timeline of all mother/infant pairs where a subject received three or more antibiotics prescriptions (eleven subjects across ten pairs). Here we see that repeated antibiotics prescriptions typically form a contiguous sequence across multiple visits, and that concurrent antibiotics use in mothers and infants is common. We also highlight the infrequent prescriptions for macrolides and sulfonamides, both of which are recommended treatments for *B. pertussis* infections. Our results indicate persistent pertussis infections in four infants (C, E, F, and H) and three mothers (D, E, and G), and suggest that pertussis infections persist during and after antibiotics use.

**Figure 4.**
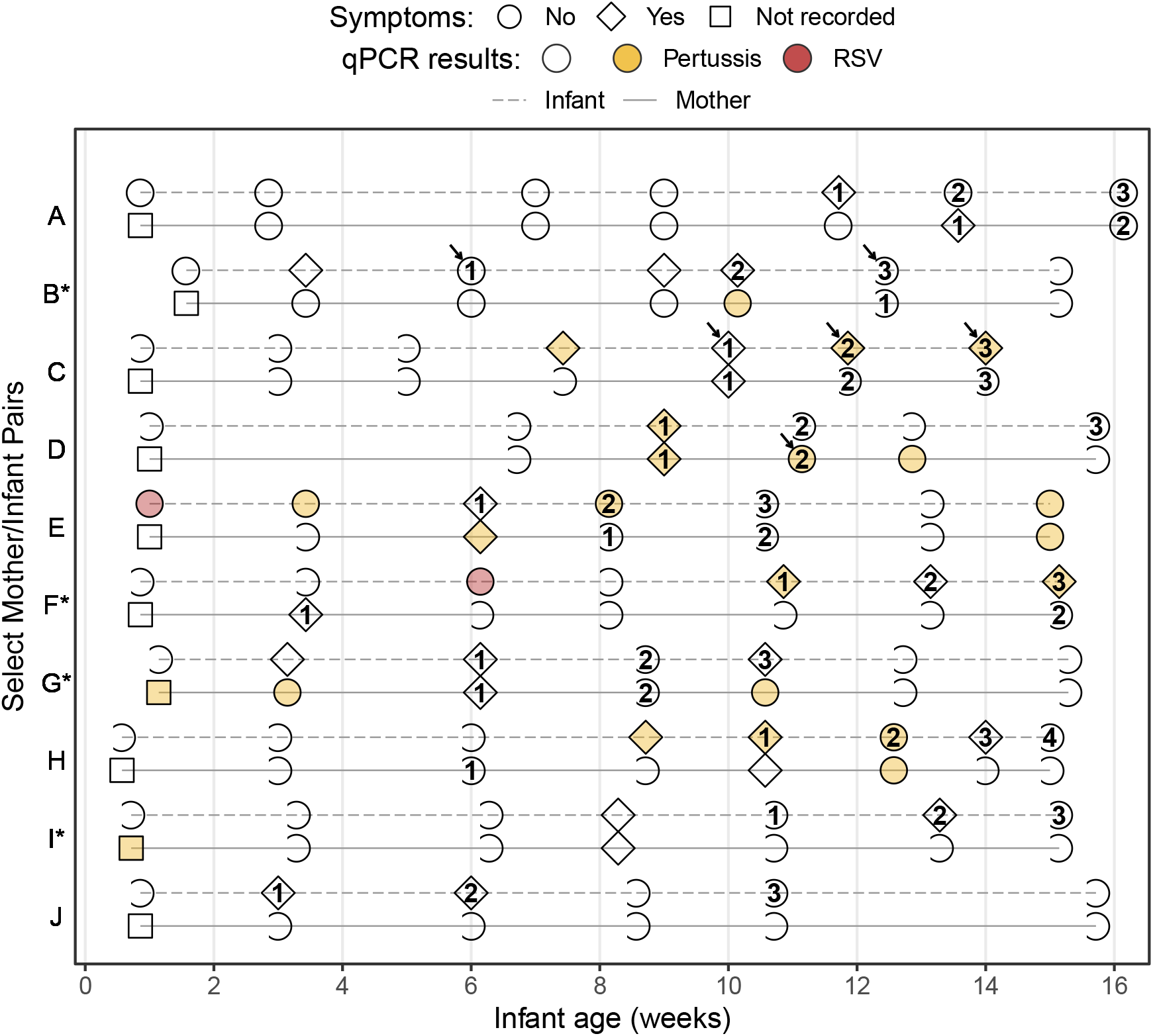
Study timeline of all mother/infant pairs where a subject was prescribed antibiotics at ≥3 visits (pairs are ordered by enrollment date, * indicates HIV+ mothers). Shape indicates presence of symptoms, color shows qPCR results, and numbers show cumulative antibiotics prescriptions. Arrows indicate prescription of macrolide (infants B and C) or sulfonamide (mother D) antibiotics. Clinical records were not collected at enrollment, but only infants without apparent disease were enrolled. Of ten infants with ≥3 antibiotics prescriptions, four had multiple pertussis detections (RR=4.9, p=0.024; infants C, E, F, and H) and two had an RSV detection (RR=8.9, p=0.028; infants E and F).

## Discussion

In this study, we leverage intensive monitoring of mother/infant pairs during routine well-child health care across newborn infants’ first four months of life to provide a fine-grained view of realized health outcomes. Through longitudinal surveillance of mother/infant pairs, we document the intensity of antibiotic use over time and quantify the relative impacts of prior and current infections on antibiotics use. Prospective surveillance also allows us to draw conclusions about the study population at large, relative to opportunistic sampling of symptomatic children and their mothers. We posit that our study population is broadly representative of risks and challenges facing at-risk urban communities worldwide, including rapid growth, limited access to health care, economic constraints on treatment options, and major gaps in public health disease surveillance [40–42].

We were surprised at the widespread presence of *B. pertussis* in both infants and mothers (8% and 9% of NP samples, resp.), albeit often at very low levels (i.e., high CT values). Yet the observed correspondence between symptoms and more intense IS481 CT values in infants highlights the clinical relevance of our findings. Overall, we found that antibiotics use was widespread, particularly in infants, and consisted mostly of penicillins and cephalosporins. Yet macrolides and sulfonamides, which can effectively treat *B. pertussis*, were only rarely prescribed. As expected, we observed a strong association between symptoms and antibiotics use, both of which more common when *B. pertussis* and RSV were present, indicating that inappropriate antibiotics were commonly prescribed to treat these two pathogens.

A noteworthy limitation of this study is the infrequent detection of RSV, which offers limited statistical power for inference. On one hand, we expected to detect RSV more frequently than *B. pertussis* in infants, as no RSV vaccine is available and almost all children contract RSV by three years of age [43–45]. On the other hand, maternal antibodies provide some protection during infants’ first 1-2 months of life [45], while RSV’s shorter duration of infection would yield fewer detections relative to pertussis at a comparable incidences. Our results also suggest that we captured the tail of an RSV outbreak in this population while the cohort size was small but increasing (due to rolling enrollment), and while enrolled infants were no more than 2 months of age.

The observational nature of this study is both a strength and a limitation. We cannot confidently ascribe causal relationships to the observed patterns, nor can we rule out the impact of unobserved, epidemiologically important covariates on our findings. However, by prospectively recording the delivery of pediatric care in a representative clinical setting, we were able to characterize drug-pathogen interactions that, due to logistical constraints and ethical considerations, would be difficult or impossible to evaluate in clinical trials. Similarly, our retrospective qPCR testing of NP samples precludes any clinical impact of test results, while our focus on population-level surveillance allows us to use more sensitive (and possibly less specific) qPCR signals for pertussis than would be appropriate for clinical disease diagnosis. Consequently, this study provides an incomplete but nuanced view of realized health care outcomes in a representative low-resource urban community.

### Public health impacts for low-resource settings

Trade-offs between over- and under-treatment pose a recurring dilemma in routine clinical care. This dilemma is particularly acute in low-resource settings, where staff, facilities, and diagnostic capacity are limited, and where treatment costs are constrained. *A priori*, we expect that public health clinicians seek to minimize ineffective use of antibiotics in order to reduce avoidable disease, side-effects, and financial costs. Yet the immediate costs of inappropriately treating, e.g., *B. pertussis* with amoxicillin, may be low relative to the risks of *failing* to treat sensitive pathogens, especially in pediatric care.

We have demonstrated the routine use of penicillins in our cohort as a non-specific (and potentially prophylactic) treatment of respiratory symptoms, particularly in infants. By pairing clinical records with molecular surveillance, our study highlights a notable gap in current IMCI guidelines regarding pertussis. Our results indicate that minimally symptomatic pertussis was frequently treated with by PHC clinicians with inappropriate non-macrolide antibiotics, and that infections commonly persisted in the absence of appropriate chemical treatment. We note that a recent IMCI update recommends the addition of an inexpensive macrolide for symptomatic children over three years of age who have completed an initial round of amoxicillin and are “not better but not worsening at time of re-assessment”, but do not directly address *B. pertussis* [11]. We also note the virtual absence of reported pertussis cases in Lusaka, despite evidence for widespread asymptomatic and minimally symptomatic pertussis in this cohort [36]. We speculate that improved diagnostic specificity and/or clinician education paired with timely, appropriate treatment of pertussis in both infants and their mothers could substantially reduce the burden of disease in this population while reducing off-target use of penicillins.

## Materials and Methods

### Study Design

This work provides a retrospective analysis of observational data collected during the Southern Africa Mother Infant Pertussis Study (SAMIPS). SAMIPS was a longitudinal cohort study in Lusaka, Zambia during 2015 that followed mother/infant pairs across infants’ first four months of life. A detailed account of study methods, including sample size considerations, eligibility criteria and enrollment, nasopharyngeal (NP) sample collection and handling, and routine infant vaccinations have been previously published [35–37]. Here we provide a brief overview.

SAMIPS sought to enroll all healthy live births in Chawama township that occurred between March and December 2015. Enrollment was conducted at the Chawama Primary Health Clinic (PHC), and mother/infant pairs were recruited during their first scheduled postpartum well-child visit at approximately 1 week of age. Mothers were provided with modest incentives to join and remain in the cohort. Note that PHC is the only government-supported clinic, and the only source of no-cost health care, in this community. Eligible infants were full-term, delivered without complications, and symptom-free at enrollment. Maternal eligibility included signed consent, Chawama residency (anticipated remaining in the community during study period), known HIV status prior to delivery, and treatment with prophylactic antiretroviral therapy at the time of delivery for HIV+ mothers.

After an initial enrollment visit, mother/infant pairs were scheduled for six routine followup clinic visits at 2-3-week intervals through approximately 14 weeks old (maximum, 18 weeks). Additional unscheduled clinic visits were initiated by study mothers for acute medical care as well as routine well-child care. As noted above, we exclude unscheduled visits from consideration here.

At each clinic visit (including enrollment), NP swab samples were obtained from mother and infant. At each visit except for enrollment, detailed records of self-reported symptoms of respiratory infection were collected by clinic staff. Pre-printed barcodes and data entry forms were used to track subjects, clinic records, and NP samples. The Xcallibre digital pen system was used to automatically record dates and times, scan barcodes, and digitize hand-written text.

For antibiotics prescriptions, the medication name and dose was recorded as free-form text, and duration in days was also recorded. For symptoms, mothers were asked if the following symptoms were present since their previous clinic visit in themselves and their infants: cough, coryza, uncontrollable coughing, whooping, posttussive vomiting, cyanosis, labored breathing, and wheezing, and (only in infants) hot/feverish, difficulty feeding, lethargy, fits or seizures, and apnea. Clinicians directly observed and documented the presence of cyanosis, paroxysmal cough, whoop, apnea, conjuctival injection, mechanical sequellae of cough, lethargy, and bronchitis or pneumonia, and (only in infants), poor suck, seizure, and chest wall in-drawing. Clinicians also measured and recorded temperature. We define fever as a temperature above 38 degrees C. Note that, while symptoms were not systematically recorded at the enrollment visit, enrollment criteria specified that infants (but not mothers) were free of apparent symptoms.

Routine childhood vaccinations were administered by regular clinic staff at appropriate clinic visits according to the official Zambian schedule (for details, see Gunning et al. [37]). All vaccinations were administered after NP samples were collected and in a separate area of the clinic in order to avoid possible cross-contamination between NP samples and vaccines.

### Laboratory Methods

NP sample DNA and RNA were extracted using the NucliSENS EasyMag system (bioMérieux, Marcy l’Etoile, France). We used quantitative PCR to test samples for human RNAse P (RNP), *B. pertussis*, and RSV. RNP is a the constitutively expressed gene that we used to assess successful sample collection, storage, DNA extraction, and lack of PCR inhibition. The *B. pertussis* assay was a singleplex TaqMan qPCR reactions targeting the IS481 insertion sequence [46], and the RSV assay used a reverse-transcriptase qPCR reaction [47]. Samples were run on 96-well qPCR plates; each plate contained approximately 46 samples (one each of IS481 and RNP), along with one positive and one negative control per plate. Each plate was run for 45 cycles, such that the minimum detectable target quantity had a CT value of 45 (lower values indicates more target). We consider assays with a CT value of 45 or less to be detecting assays; all others were considered non-detecting (N.D.).

All primers and probes were purchased from Life Sciences Solutions (a subsidiary of ThermoFisher Scientific Inc). Most plates were run on an ABI 7500 thermocycler (ThermoFisher Scientific Inc, Waltham, MA). Starting in 2019, some plates were run on a QuantStudio5 thermocycler (ThermoFisher Scientific Inc, Waltham, MA). A subset analysis of samples run in parallel on both machines showed minimal systematic variation between machines, and we do not distinguish between machines here.

### Statistical Analysis

We focus here on the 1,320 mother/infant pairs who have ≥ 4 NP samples per subject, which is the same analysis set as in Gill et al. [36]. We have excluded unscheduled clinic visits and limit our analysis here to enrollment and scheduled clinic visits in order to avoid the potentially confounding effects of self-initiated health care and more clearly focus on routine surveillance.

In order to categorize prescriptions by antibiotic mode of action, we manually inspected free-form text, and then developed a set of regular expressions that accounted for common abbreviations and optical character recognition errors. Prescription could include more than one medication and corresponding category.

Our temporal analysis used a set of generalized additive models (GAMs) to estimate the relative frequency of pathogen detection throughout the study (one model per pathogen, Figure 2A-B). Our GAMs use a binomial link function, and were smoothed by calendar date using cubic regression splines with shrinkage. For inference, we used a set of generalized linear models (GLMs) to estimate the per-visit probability of A) respiratory symptoms (Table 3) and, B) antibiotics prescription (Table 4), with separate models for mothers and infants. The GLMs estimating antibiotics prescriptions used observations from both the current and previous visit as covariates. Initial enrollment visits were thus excluded from the antibiotics GLMs, as antibiotics were not recorded at enrollment.

All analysis was conducted in R version 3.5.2 [48]. We used a Mann-Whitney test to compare IS481 CT values between subjects with and without symptoms (Figure 3. We used the Wald method (unconditional maximum likelihood estimation) in the epitools package to compute relative risk and corresponding p-values (Figure 4).

## Data Availability

Data and code will be provided at OSF at time of publication.

https://osf.io/tnkyu/

## Acknowledgments

We wish to thank Rachel Pieciak, William MacLeod, Magdalene Mwale, Arash Saeidpour, Deven Gokhale, and Tobias Brett for their helpful comments. We also wish to thank the Lusaka lab team who generated the results for this analysis: Caitriona Murphy; Ruth Nkazwe; Chilufya Chikoti; and Baron Yankonde. The lab testing and subsequent analyses for this paper were supported by a grant from the National Institutes of Health/National Institute of Allergies and Infectious Diseases (R01AI133080). Funding for the initial SAMIPS study that allowed us to create the sample library itself was through the generous support of the Bill and Melinda Gates Foundation (OPP1105094).

